# APOE-specific Cognitive Effects of Levetiracetam in Mid-Age Adults

**DOI:** 10.64898/2026.05.14.26352135

**Authors:** C. Lancaster, N. G. Dowell, G. Tertikas, A. Bakker, C. Bird, N. Tabet, J. Rusted

## Abstract

**Background:** Low-dose levetiracetam is under investigation as a potential treatment for slowing Alzheimer’s Disease progression. This study tests whether levetiracetam enhances executive function in mid-age adults, and whether drug effects differ by Apolipoprotein e4 (APOE4+) genetic risk status.

**Methods:** Fifty-eight adults (aged 45-65 years; 27 APOE33; 31 APOE4+) participated in a double-blind, placebo-controlled study of low-dose levetiracetam (125mg bidaily for two-weeks). At the end of each treatment phase, participants completed a switch-inhibition task.

**Results:** Mid-age APOE4+ carriers were significantly slower and showed a greater cost of increasing executive demand than APOE33 individuals. Response times were quicker under levetiracetam, with increased benefits reported in APOE33 individuals, at younger ages, and in individuals with reduced levels of plasma-based biomarkers. Levetiracetam selectively benefitted accuracy in APOE33 individuals.

**Conclusion:** Low-dose levetiracetam enhances executive function in midlife, particularly in individuals at lower risk of Alzheimer’s Disease based on age, APOE4 genotype, and proxies of neuropathology.

## 1. Introduction

The development of treatments which slow or halt the progression of Alzheimer’s Disease (AD) has largely focussed on clinical trials targeting amyloid-β and tau pathologies[1]. The complex aetiology of AD however, encompassing genetic risk factors such as Apolipoprotein ε4 (APOE4) and including both disease-specific and non-disease-specific pathways, underlines the need for strategies which target multiple contributors to cognitive decline[2:4]. Acknowledging the extended preclinical phase of AD[5], clinical trials increasingly seek to alter disease onset from earlier in the lifespan, focusing on high-risk individuals in the mild cognitive impairment (MCI) phase of the disease and even earlier[2]. This study provides novel exploration of whether low dose levetiracetam benefits cognition in mid-life carriers of Apolipoprotein e4 (APOE4+), the most well-established genetic risk variant for sporadic Alzheimer’s Disease [6].

Neuronal hyperactivity, particularly in the hippocampus, is well established in the early stages of AD[7:9]. Indeed, evidence from preclinical and human studies suggest neuronal hyperactivity drives amyloid-β and tau accumulation, triggering widespread network dysfunction, neurodegeneration, and associated cognitive decline[10:14]. Low-dose levetiracetam - a commonly prescribed anti-seizure medication-, is under investigation as a potential treatment to slow progression from MCI to AD, hypothesised to act on such signatures of cerebral hyperactivation[7,15]. Importantly administration of levetiracetam, but not other anti-seizure medications, is reported to modify patterns of cortical (including hippocampus) hyperactivity in human amyloid precursor protein (hAPP) transgenic mice [16]. Furthermore, acute administration of low-dose levetiracetam downregulates hippocampal hyperactivity in individuals with a diagnosis of MCI[7,17], a brain region critically involved in the clinical progression of AD.

Cognitively, preclinical evidence indicates levetiracetam is associated with improvements in spatial and associative-memory in aged mice[18,19] and hAPP mice[16,20,21], though not consistently[19]. Meta-analyses of the small number of human randomised control-trials (RCTs) exploring the cognitive effects[22,23] of levetiracetam provide preliminary support for domain-specific benefits of this drug in executive function and visuospatial processing[24,25], however, conclusions did not account for possible variable effects of drug dose[17]. There is limited support to date that longer-term administration of levetiracetam is associated with significant slowing of global cognitive symptoms in individuals with MCI[26]. When differentiating by APOE4 genotype however, only non-carriers showed significant slowing of entorhinal cortex atrophy after 18 months of treatment, correlated with reduced change in cognitive function, and lower levels of plasma biomarkers of neuronal damage and inflammation[27]. It may be that the beneficial cognitive effects of low-dose levetiracetam may be enhanced if administered earlier in the lifespan, for example in mid-life, to coincide with emerging trajectories of increased AD pathology.

APOE4+ carriers are an important target for preventative interventions, especially given evidence anti-amyloid therapies are both less effective and safe for use in this group[28]. APOE4+ individuals may benefit from prophylactic use of low-dose levetiracetam as patterns of aberrant brain activity are reported in cognitively healthy APOE4+ across the lifespan (e.g.,[29:34], but see also[35,36]). This is corrobotated by preclinical evidence linking APOE4+ to a toxic excitatory phenotype from youth[37]. Although APOE4+ show equivalent performance on neuropsychological test batteries in mid-life[38], more nuanced behavioural paradigms - designed for use in clinically healthy adults, expose subtle behavioural deficits emerging by mid-life in APOE4+[e.g., 39:44], particularly on tests of executive function. For example, consistent with studies reporting very early sensitivity of Stroop-switch paradigms to AD[45,46], mid-age APOE4+ are reported to show increased errors on this paradigm in mid-life[40].

Here, we tested if low-dose levetiracetam enhanced cognitive performance on a comparable executive challenge task in mid-adults differentiated by APOE4 genotype, utilising a placebo-controlled crossover design. Under placebo, mid-age APOE4 carriers were predicted to show executive deficits relative to an APOE33 control group, with differences anticipated to be greatest on trials with high executive load. Low dose levetiracetam was expected to act as a cognitive enhancer, with benefits greatest in at-risk mid-age APOE4 individuals. Further exploratory analyses tested whether potential cognitive effects of low-dose levetiracetam are influenced by plasma-biomarkers of neuronal health, including proxies of AD pathologies (amyloid-β-42 (Aβ_42_), amyloid-β-40 (Aβ_40_), phosphorylated tau-181), neurodegeneration (neurofilament light (NfL)), and astroglial activation (glial fibrillary acidic protein (GFAP)).

## 2. Methods

### 2.1 Study design

A double-blind, placebo-controlled, within-subject crossover design was used to test the cognitive effects of low-dose levetiracetam in 60 middle-aged (45-65 years) adults differentiated by APOE4 genotype. Following the procedure of Bakker and colleagues[7,17], 125mg of levetiracetam or a visually matched placebo was administered bidaily (BID; morning, evening) to participants for 14-days, followed by a study break lasting a minimum of 4-weeks. Following this, participants received the alternate treatment (125mg levetiracetam or placebo BID) for a further 14-days. Treatment order (levetiracetam first, placebo first) was randomised for each participant by a third-party researcher, ensuring an equivalent number of APOE4+ and APOE33 individuals per protocol. Compliance, mood, and sleep were recorded daily during the treatment phase using the Mezurio smartphone app[47,48]. At the end of each treatment phase, participants completed cognitive and neuroimaging assessments, plus provided a blood sample. All adverse events were recorded during the study period; moderate to severe adverse events that could possibly be linked to the study drug resulted in participant unblinding to drug condition and withdrawal from the study.

The study was approved by the Brighton & Sussex Medical School Research Ethics Committee (ER/CLL62/1) and was completed in accordance with the ethical standards outlined in the 1964 Declaration of Helsinki (and its later amendments). Levetiracetam was used in this project as a pharmacological manipulation to advance understanding of the relationship between fMRI BOLD activation and cognitive processes such as executive function; specifically, how this relationship may differ as a consequence of APOE genotype. As we are not testing its clinical efficacy or safety profile, it was not deemed a clinical trial by the UK Medical and Healthcare Products Regulatory Agency.

### 2.2 Participants

A total of 305 volunteers were recruited to join an APOE database through advertisement in the local community (including the University) and the Join Dementia Research platform (UK-based). In addition, invitation to participate was extended to individuals in the existing APOE research database at the University of Sussex. For inclusion, participants were required to be non-Hispanic white, English-speaking, and have access to an Android or Apple smartphone. Participation was limited to individuals from a non-Hispanic white background as the risk associated with APOE4 differs by race[49] and sample size constraints prohibited stratifying genotype effects by ethnicity.

Exclusion criteria: past or pre-existing diagnosis of cognitive impairment (e.g., MCI, AD), any psychiatric or neurological condition including epilepsy, a history of seizures or use of anti-seizure medications, current use of neuroactive medications (anxiolytics, narcotics, anti-depressants, anti-psychotics, opiates, antihistamines with anticholinergic properties), or contraindications for MRI. In addition, volunteers entering the study were excluded if they had a known allergy to levetiracetam or previous significant sensitivity to medication, a history of anaphylactic shock, were under investigation for any suspected liver or kidney disorders or had a current diagnosis of significant medical illness (e.g., chronic renal or liver failure, heart disease). Mid-age females were screened for pregnancy or current breast-feeding.

APOE genotyping followed Human Tissue Authority (HTA) procedures. Specifically, DNA was collected with a buccal swab, using an Isohelix SK1 kit. Samples were analysed to determine APOE gene variant by LGC Genomics (Hertfordshire, www.lgcgroup.com/genomics). A fluorescence-based competitive allele-specific polymerase chain reaction determined the presence of three major APOE alleles (e2, e3, and e4) based on two APOE single nucleotide polymorphisms (SNPs) (rs429358, rd7412). Double-blind procedures were followed through-out; APOE genotype was not disclosed to the participant during or after completion of the study.

Following screening for APOE genotype, an independent third-party researcher (randomisation lead) selected participants for the main study to ensure ~ 50% APOE4+ (APOE34, APOE44) in each age-group, and that sex and age remained balanced across genotype groups. APOE e2 were excluded at this stage due to the reported protective effects of this less common variant against AD[50]. Demographic characteristics of the final sample are shown in Table 1.

**Table 1.** Demographics presented by Genotype Group. Values represent mean ± sd unless stated otherwise. * denotes significant differences by APOE4 status or APOE4 gene dose.

| APOE33 | APOE4+ | APOE34 | APOE44 |
| --- | --- | --- | --- |

| *n* | 27 | 31 | 23 | 8 |
| --- | --- | --- | --- | --- |
| Age (Years) | 54.11 ± 4.91* | 56.52 ± 5.06 | 55.57 ± 5.39* | 59.25 ± 2.60* |
| Sex (%F) | 70.37 | 77.42 | 82.61 | 62.5 |
| Education (Years) | 17.37 ± 3.31 | 18.26 ± 4.00 | 18.13 ± 3.81 | 18.63 ± 4.78 |
| Full-scale Premorbid IQ | 119.55 ± 4.09* | 117.64 ± 4.55 | 118.36 ± 4.77* | 115.55 ± 3.23* |
| MoCA (/30) | 27.81 ± 1.73 | 27.26 ± 1.84 | 27.43 ± 1.70 | 26.75 ± 2.25 |
| Plasma levetiracetam concentration(mg/L) | 3.53 ± .84 | 3.19 ± 1.01 | 3.37 ± 1.07 | 2.51 ± .85 |
| Aβ<sub>42:40</sub> (pg/mL) | .07 ± .01 | .06 ± .01 | .06 ± .01 | .06 ± .01 |
| pTau181(pg/mL) | 20.16 ± 8.34 | 24.02 ± 14.32 | 24.90 ± 16.09 | 21.52 ± 7.68 |
| GFAP (pg/mL) | 74.70 ± 23.24 | 121.46 ± 73.28 | 113.80 ± 73.13 | 144.41 ± 74.29 |
| NfL (pg/mL) | 8.68 ± 2.46 | 13.17 ± 5.51 | 13.27 ± 6.24 | 12.89 ± 2.56 |

Abbreviations: Montreal Cognitive Assessment (MoCA), Plasma Aβ<sub>42:40</sub>, phosphorylated-tau-181
(pTau181), Neurofilament light (NfL), Glial Fibrillary Acidic Protein (GFAP).

### 2.3 Plasma-based analyses

Plasma was isolated from EDTA blood samples collected from participants at the end of each treatment phase. Levetiracetam concentration (mg/L) was extracted by the Chalfont Centre for Epilepsy using an assay with a 1mg/L lowest limit of quantification. Plasma Aβ_40_, Aβ_42 (_subsequently converted to Aβ_42:40_), phosphorylated-tau-181 (p-tau-181), Neurofilament light (NfL), and Glial Fibrillary Acidic Protein (GFAP) concentrations were extracted by the UK Dementia Research Institute Biomarker Factory (UCL). Higher values of p-tau-181, NfL, and GFAP proxy poorer neurological health, whereas a lower Aβ_42:40_ is predictive of greater pathology.

### 2.4 Cognitive Assessment

All participants completed the Montreal Cognitive Assessment (MoCA) and National Adult Reading Test (NART[51]) - total number of correct words on the NART was used to estimate pre-morbid full-scale IQ.

#### 2.4.1. Switch-Inhibition Task

On each trial (*n*=72) of the computerised Switch-Inhibition Task (Figure 1), participants were presented with a single stimulus; either a rectangle, left-pointing block arrow or right-pointing block arrow, coloured orange or blue dependent on trial-type (‘location’, ‘direction’ respectively). The stimulus could be presented in three locations: in the centre, on the left, or on the right-hand side of the screen. Prior to each trial, participants were presented with a ‘pre-cue’ for 1500ms, which would tell them which rule to apply to the subsequent trial. If they were presented with ‘LOCATION’ (n=38, orange text), they needed to respond whether the stimulus for the associated trial appeared on the left or right of the screen. If they were presented with ‘DIRECTION’ (n=38, blue text), they needed to respond whether the stimulus was a left- or right-pointing arrow. Participants were instructed to respond as quickly and as accurately as possible, pressing ‘q’ for left, and ‘p’ for right (corresponding to the hand and side of keyboard). Stimuli stayed on screen until a response was made.

**Figure 1.**
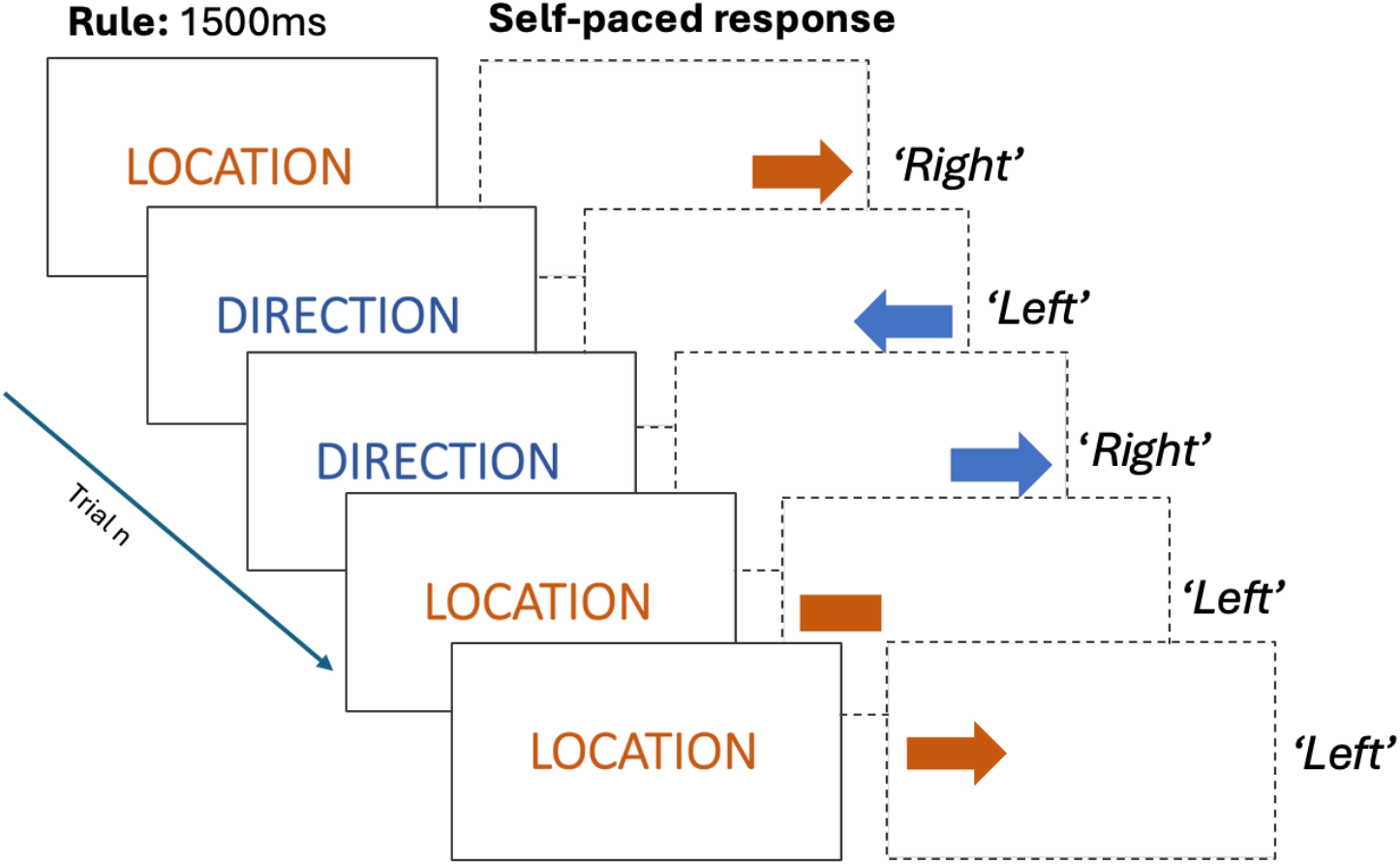
Visual representation of the switch-inhibition task illustrating 5 trials; the pre-cue ‘rule’ is presented for 1500ms followed by either a congruent, incongruent or neutral trial. The correct response for each trial is shown in *italics*.

All participants received the same, fixed order of stimuli. Switches between location and direction trials occured with an unpredictable, pseudorandom lag of 0 to 5 trials. 19 location and 19 direction trials were designated switch trials, meaning the rule changed from the proceeding trial. Trials differed in the congruency of stimulus shape – location pairings. Congruent trials include a directional shape (e.g., left-pointing arrow, right-pointing arrow) presented on the corresponding side of the screen (e.g., left-hand, right-hand respectively). Incongruent trials include a directional shape (e.g., left-pointing arrow, right-pointing arrow) presented on the opposite side of the screen (e.g., right-hand, left-hand respectively). Neutral trials include either a rectangle on the left or right-hand side of the screen, or a directional cue in the centre of the screen. There were 26 congruent, 31 incongruent and 19 neutral trials. Note, some trials included switch and inhibitory demands, exerting a dual-executive load.

### 2.5. Statistical Analyses

Hypotheses, experimental design, and statistical analyses were pre-registered on the Open Science Framework (OSF; https://doi.org/10.17605/OSF.IO/H39RP); deviations from pre-registered analyses are flagged in text.

No individuals met pre-specified grounds for exclusion, namely scoring at or below chance accuracy (50%) in response to either pre-cue (location or direction). Of note, 13 individuals (5 APOE33, 5 APOE34, 3 APOE44) scored less or equal to 26 on the MoCA (range: 22 – 30 in current study); models including standardised MoCA scores are reported in Supplementary Materials. Mid-age participants with missing data at one timepoint (*n*=4) were still included in all generalised linear mixed-effect models (GLMMs).

Group differences (APOE4+, APOE33) in demographic characteristics, plasma levetiracetam concentration, and biomarkers were probed using chi-squared tests (categorical), *t*-tests (parametric) or Mann-Whitney U tests (non-parametric) dependent on data type. As sufficient APOE44 individuals were recruited for exploratory tests of gene-dose effects, one-way analysis of variance (ANOVA; parametric) and Kruskal-Wallis (non-parametric) tests were used to screen group-differences by APOE4 allele number.

#### 2.5.1 Switch-Inhibition Performance

Switch-inhibition performance was analysed using GLMMs computed by maximum-pseudolikelihood estimation (R Package: lme4, version1 −1.32). Frequentist statistics are reported for main effects and interaction terms, plus pairwise comparisons of interest (*p*-values adjusted using Bonferroni method for multiple comparisons). Anonymised trial-level datasets are uploaded to the OSF (https://osf.io/2h3m9/files/rywtu).

Neutral trials were excluded from the reported GLMMs because no beneficial effect of congruency relative to neutral trials was observed (models including neutral trials are reported in the Supplementary Materials). Model fit, assessed using the Akaike information criterion (AIC), improved following their removal. Additional exploratory analyses reported in the Supplementary Materials include APOE gene dose (ε33 = 0, ε34 = 1, ε44 = 2) as a linear predictor of task performance, and include sex (0 = male, 1 = female) and z-standardised MoCA score as covariates.

##### 2.5.1.1 Response time

Trial response time (RT; seconds; correct trials only) was analysed using GLMMs with a gamma distribution and log link. Model 1 included the following fixed predictors: trial congruency (congruent = 0, incongruent = 1), trial switch (no switch = 0, switch = 1), APOE4 status (ε33 = 0, APOE4 carrier = 1), drug condition (placebo = 0, levetiracetam = 1), and age (years, z-standardised), together with all possible fixed-effect interactions. Random intercepts were included for participants.

Individual plasma biomarkers were entered into 4 separate exploratory RT models (corrected α=.05/4) as linear, z-standardised predictors. Prior to standardisation, biomarker values exceeding ±2 standard deviations from the sample mean were excluded.

##### 2.5.1.2 Accuracy

Accuracy (correct vs. incorrect responses) was analysed using GLMMs with a binomial distribution and logit link, such that parameter estimates reflect the probability of a correct response on each trial. Model 2 included the same predictors as Model 1; however, as age (restricted range: 45–64 years) did not significantly predict accuracy and inclusion of this fixed effect reduced model fit, this variable was not included in the reported model. Including age as a covariate did not qualitatively affect model outcomes. Individual plasma biomarkers were entered into separate exploratory models of response accuracy as linear, z-standardised predictors.

##### 2.5.1.3 Accuracy Cost

The cost of trial congruency (proportion accuracy: congruent – incongruent trials), trial switch (proportion accuracy: no-switch – switch trials), and dual executive demand (proportion accuracy: congruent no-switch – incongruent switch trials) was calculated for each participant, for each timepoint (placebo, levetiracetam). Cost metrics were subject to permutation-based analysis of generalised variance analysis (ANOVA) (R Package: permuco), including drug condition (within-subject: placebo, levetiracetam) and APOE4 status (between-subject: APOE33, APOE4+), with participant specified as a random effect. P-values were derived from 5,000 permutations using the Kherad-Pajouh and Renaud method. ANOVAs were repeated including APOE4 gene dose as an exploratory predictor (see Supplementary Materials).

## Results

APOE4+ individuals did not significantly differ from their APOE33 counterparts in sex distribution, education level or estimated full-scale IQ (p>.05), however, the APOE4+ group trended to be older (W=298, p=.061). Plasma biomarker outcomes are available for 50 participants (Table 1; 22 APOE33; 21 APOE34; 7 APOE44). APOE4+ individuals had a significantly lower Aβ42:40 ratio than non-carriers (*t*(48)=2.27, *p*=.028), plus significantly higher GFAP(*t*(33.65)=-3.18, *p*=.003) and NfL(*t*(48)=-3.55, *p*<.001). pTau-181 did not significantly differ between genotype groups (*t*(46)=-.60, *p=*.504).

### Levetiracetam intervention

Three participants withdrew from the active treatment arm of the study due to mild-moderate adverse events probably linked to levetiracetam usage (light-headedness (*n*=2); rash (*n*=1)). Mean self-reported compliance with the treatment schedule was 97.00 ± 7.38% (range 69% - 100%), including two participants who took a ‘drug holiday’ due to unrelated illness. As daily questionnaire responses ranged from 6–28 (maximum available; mean=24.11 ± 3.59), circulating Levetiracetam plasma-levels are reported in Table 1. There was no significant difference by APOE4 carrier status (*t*(43.63)=1.22, *p*=.228) or gene dose (*F*(2, 45)=2.48, *p*=.100).

Descriptive statistics showing task performance by APOE4 status are shown in Supplementary Table 1.

### Effect of e4 Status and Levetiracetam on RT

Model 1 (5974 observations; AIC = 83330; Figure 2) reports a significant main effect of trial incongruency (*t*(5940)=12.32, *p*<.001) and switch (*t*(5940)=12.22, *p*<.001) on estimated RT. As expected, longer RTs are reported for incongruent trials and switch trials. Main effects of drug condition (*t*(5904)=-4.03, *p*<.001), APOE e4 status (*t*(5904)=2.24, *p*=.025) and standardised age (years; *t*(5904)=3.17, *p*=.002) were all significant. RTs were quicker under levetiracetam and in the APOE33 control group. In addition, with increasing age, RTs increased.

**Figure 2.**
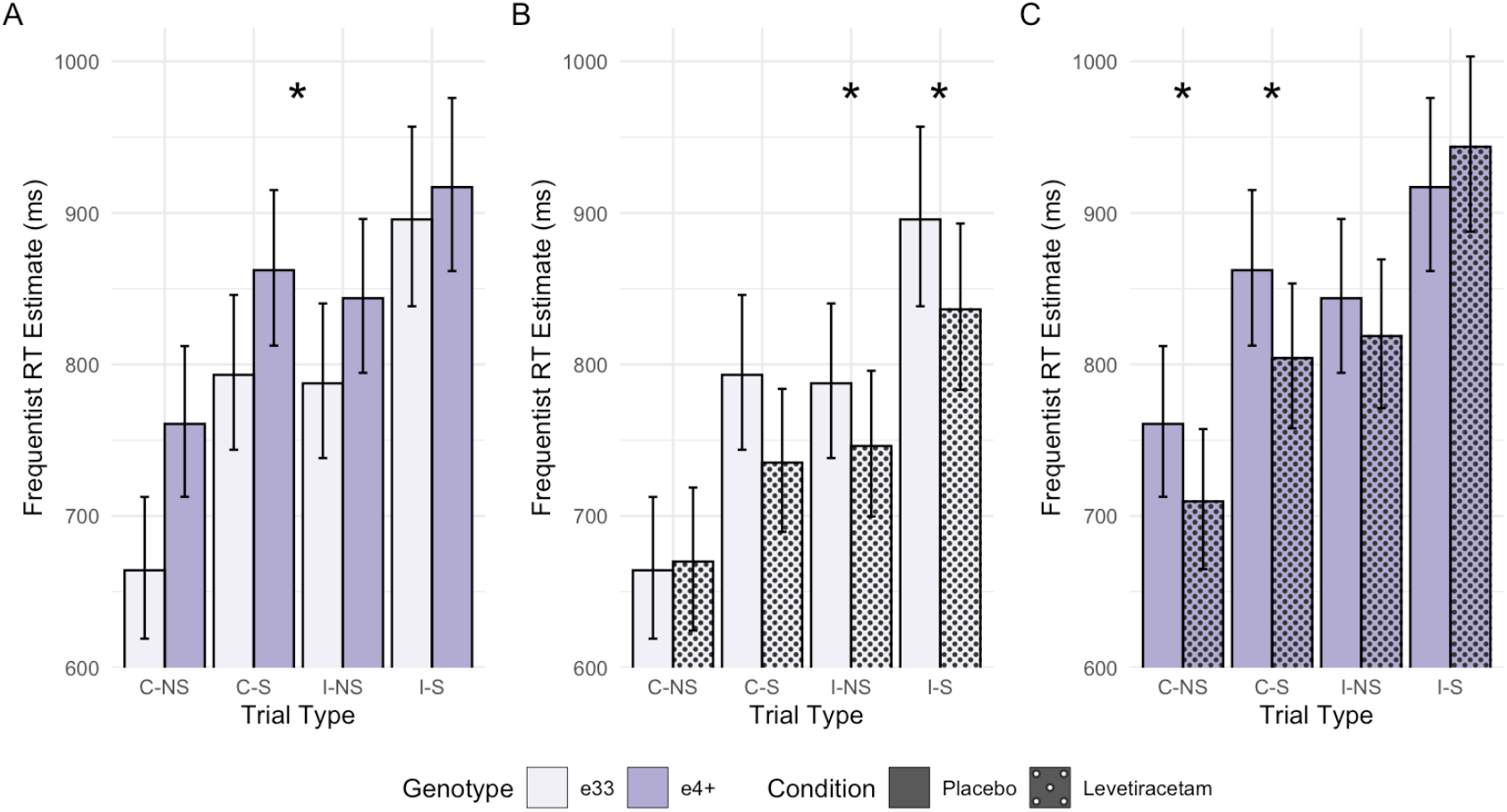
Frequentist model estimates of response time (RT) for correct trials. Estimates are shown at the mean age of the sample, grouped by trial-type, with error bars representing 95% confidence limits. **A)** APOE4 carriers are slower across trial-types under placebo (*p=*.025). **B)** Effect of Levetiracetam in APOE33 individuals, with significant beneficial effect for incongruent trials only. **C)** Effect of Levetiracetam in APOE4+ individuals, with significant beneficial effect for congruent trials only. **Note**. Trials are grouped by congruency (congruent (C), incongruent (I)) and switch (no-switch (NS), switch (S)). * denotes significant effect (*p*<.05, corrected for multiple comparisons).

The 2-way interaction between drug condition and age was significant (*t*(5904)=2.65, *p*=.008); the benefit of levetiracetam for response speed was larger in younger participants, with significant drug effects observed at the mean sample age (54.11 years; *p*<.001) and below (*p*<.001) (Figure 3).

**Figure 3.**
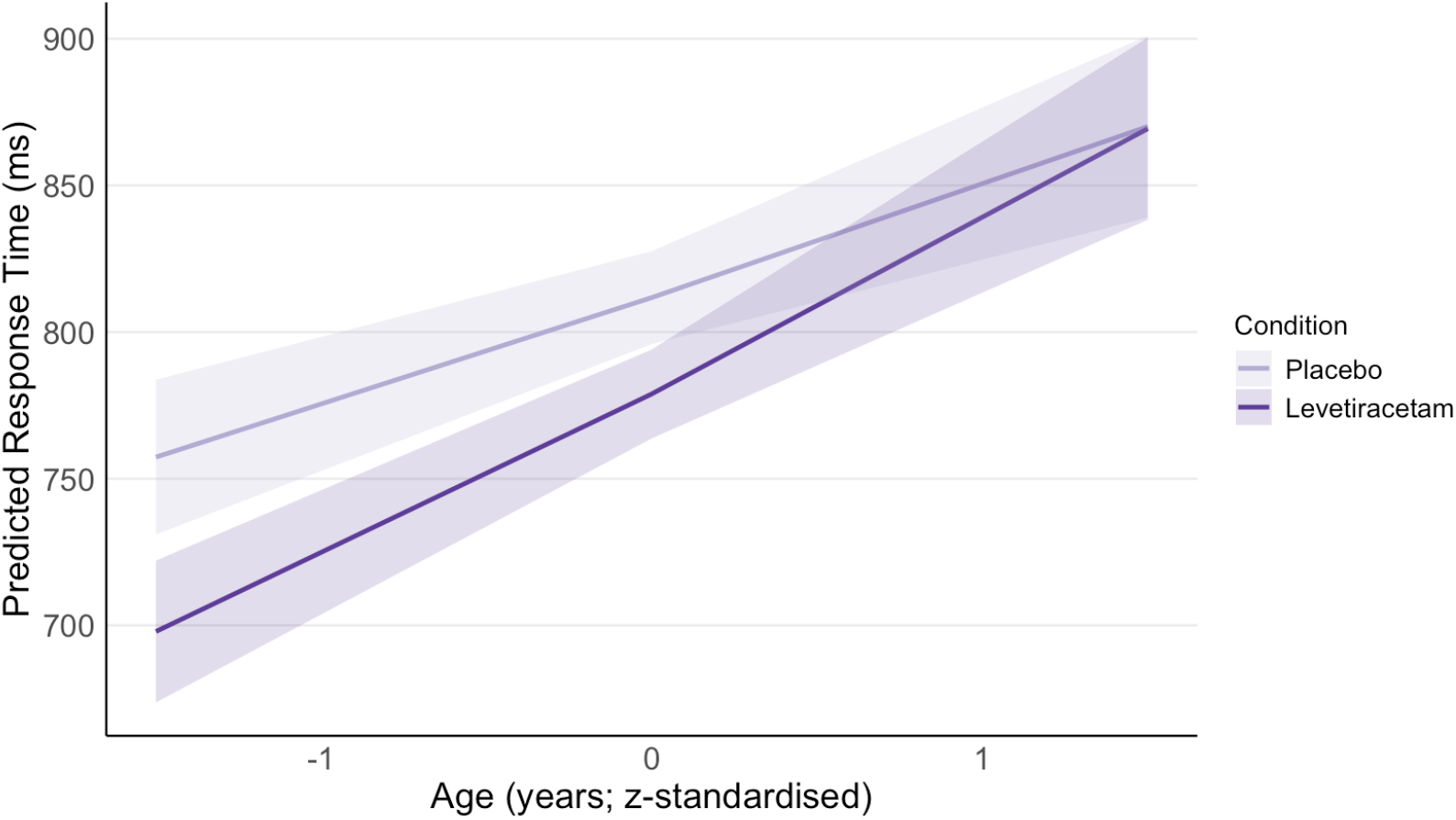
The interaction between age (years; *z-*standardised) and drug condition (placebo, low-dose levetiracetam) on frequentist model estimates of response time.

The 3-way interaction between drug condition, APOE4 status, and trial congruency was significant (*t*(5940)=2.39, *p*=.017). Post-hoc analyses (α=.006) tested if the effect of levetiracetam differed by trial congruency in each genotype group. In APOE33 individuals, beneficial effects of levetiracetam only reached significance for incongruent trials (*p*=.002), whereas in APOE4+, levetiracetam enhanced RT only for congruent trials (*p*<.001). APOE4+ were significantly slower than APOE33 individuals for congruent trials under placebo (*p*=.009) and incongruent trials under levetiracetam (*p*=.009).

### Exploratory analyses: Plasma Biomarkers

The addition of individual plasma-biomarkers as predictors in RT models improved model fit relative to Model 1 (corrected α=.013). There were no main effects for individuals plasma biomarkers on RT across models (*p*>.013), however significant interaction terms (Figure 4) are described below.

**Figure 4.**
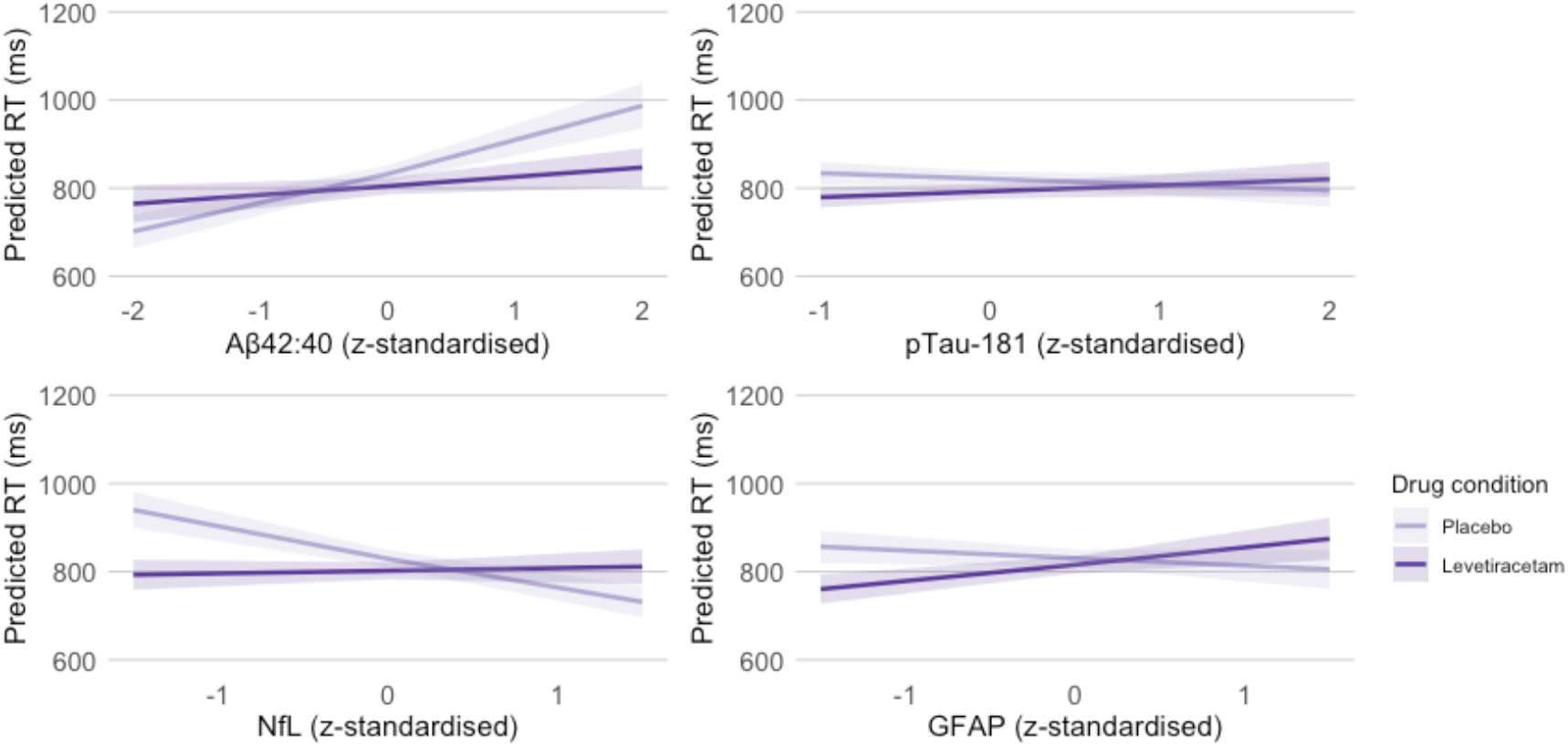
Frequentist model estimates of response time (RT, correct trials) showing the interaction between plasma biomarkers of neurodegeneration and drug condition.

There was a significant Aβ42:40 x drug condition interaction (*t(*5027)=-5.08, *p*<.001); levetiracetam enhanced RT at mean and higher Aβ42:40 (*p*<.005, α=.010); conversely, leviteracetam was associated with slower RTs for participants with lower Aβ_42:40_ ratios (*p*=.001). Unexpectedly, lower Aβ42:40 was significantly associated with faster RTs under placebo only (*p*<.001).

There was a significant pTau-181 x condition interaction, *t*(4971)=3.00, *p*=.003. Post-hoc analyses (α=.008) report a significant beneficial effect of levetiracetam on RTs at mean and below levels of pTau-181 only (*p≤* .001). There was no significant relationship between pTau-181 and RT under placebo or levetiracetam (*p*>.008).

There was a significant NfL x condition interaction, (*t*(4815)=6.96, *p*<.001), with increasing levels of NfL in plasma associated with significantly slower RTs under placebo (*p*=.001, α=.007) but not levetiracetam (*p*=.768). A significant beneficial effect of levetiracetam on RTs was observed in participants with below mean levels of NfL (*p*<.001); as levels of NfL plasma increased above the sample mean, effects reversed so that levetiracetam was associated with significantly slower RTs than placebo (*p*<.001). In addition, the relationship between trial congruency and APOE4+ was modified by NfL, *t*(4815)=-2.54, *p*=.011, however, post-hoc effects did not survive multiple comparison correction.

The interaction between GFAP x condition x APOE4 is significant (*t*(4816)= −5.35, *p*<.001), plus lower-level 2-way term (GFAP x condition, *t*(4816)= 4.78, *p*<.001). In APOE33 individuals, the effect of levetiracetam on RT is moderated by GFAP level, such that individuals with below-mean GFAP levels, levetiracetam enhances RTs (*p*<.001, α=.003). However, as GFAP levels increase above the sample mean, the drug increases RTs (*p*<.001). In APOE4+ individuals, levetiracetam reduces RTs, trending towards significance only when *z-*standardised GFAP equals 0 (sample mean, *p*=.008) and 1.5 (*p*=.009). In addition, the relationship between trial congruency and APOE4+ was modified by GFAP (*t*=-2.63, *p*=.008), however, differences did not survive multiple comparison correction.

### Effect of e4 Status and Levetiracetam on Accuracy

Model 2 (6385 observations; AIC=2625; Figure 5) reports a significant main effect of trial congruency (*t*(6368)=-10.61, *p*<.001) and switch (*t*(6368)=-2.89, *p*=.004) on probability of correct response. As expected, the probability of correct response was lower for incongruent trials and for trials preceded by a rule switch. Main effects of APOE4 status (*p*=.541) and drug condition (p=.652) were non-significant.

**Figure 5.**
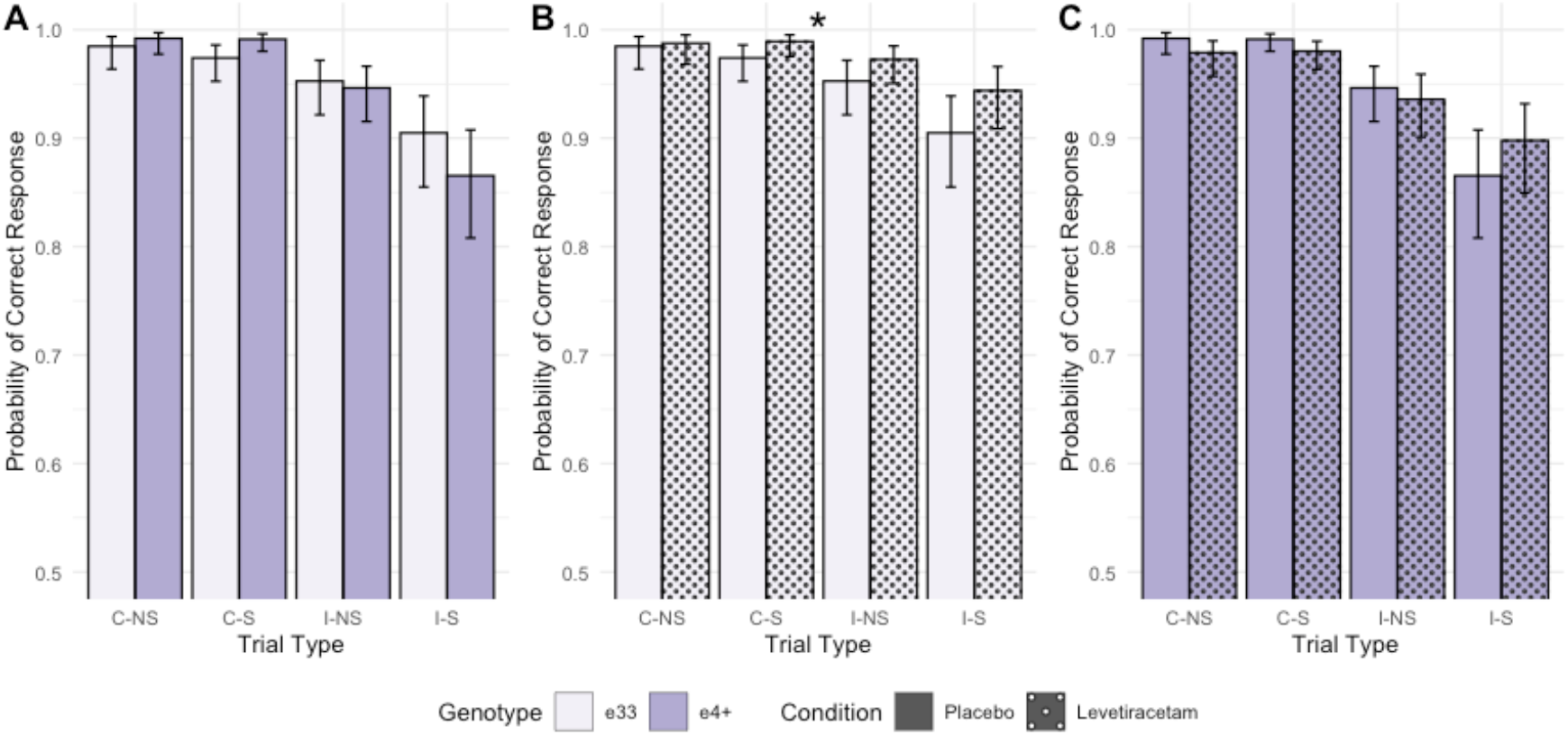
Frequentist model estimates of probability of correct response. Estimates are shown grouped by trial-type, with error bars representing 95% confidence limits. **A)** APOE4 carriers shwo greater cost of trial incongruency under placebo. **B)** Effect of Levetiracetam in APOE33 individuals is significant across trial types. **C)** Non-significant effect of Levetiracetam in APOE4+ individuals. **Note**. Trials are grouped by congruency (congruent (C), incongruent (I)) and switch (no-switch (NS), switch (S)). * denotes significant effect (*p*<.05, corrected for multiple comparisons).

The congruency x switch interaction was significant (*t*(6368)=-2.17, *p*=.030). Post-hoc comparisons (α=.013) indicate there is a significant switch-cost for incongruent (*p*<.001) but not congruent trials (*p*=.688); the effect of trial incongruency is significant for both no-switch (*z=*5.36, *p*<.001) and switch trials (*z=*10.48, *p*<.001), with a larger effect under switch conditions.

There is a significant congruency by APOE4 interaction (*t*(6368)=-2.24, *p*=.025). The cost of trial incongruency, whilst significant in both groups, is greater for APOE4 (*z=*9.23, *p*<.001, α=.013) than APOE33 individuals (*z=*5.84, *p*<.001). In addition, whilst the genotype difference does not reach significance for congruent trials (*p=*.677), it approaches significance for incongruent conditions (*p*=.068), with APOE4+ individuals performing worse. There is also a significant drug condition by APOE4 interaction (*t*(6368)=-3.28, *p*=.001). Post-hoc analyses (a=.013) report a significantly higher probability of correct response under levetiracetam in APOE33 individuals (p=.010), with this group trending to outperform APOE4+ individuals under levetiracetam (*p*=.041). Estimated probability of correct response is lower in APOE4+ carriers under levetiracetam relative to placebo (*p*=.041), but this effect does not survive multiple comparisons.

Exploratory analyses including plasma biomarkers of neuronal health as additional predictors in Model 2 revealed no significant main effects of Ab42:40, p-tau181, GFAP, and NfL on probability of correct response (*p*>.05). Further consideration of biomarker interactions with trial, genotype, or drug conditions did not improve model fit.

### Accuracy cost metrics

Secondary analyses of the cost to performance accuracy of increasing executive load (see Table 2 for descriptive statistics) revealed APOE4+ individuals (*M=*.09, *SD=*.10) showed a significantly larger cost of trial incongruency relative to their APOE33 counterparts (*M=*.05, *SD=*.07, *F*(1,56)=5.06, *p*=.030, however, both the main effect of drug condition, and APOE4 status x condition interaction were both non-significant (*p*>.05). There was no significant genotype difference in switch cost (*p*>.05), but levetiracetam (*M=*.02, *SD=*.04) was associated with a trend-level reduction in switch cost (*F*(1, 56)=3.31, *p*=.071) relative to placebo (*M=*.03, *SD=*.04). The APOE4 status by drug condition interaction term was non-significant (*p*>.05). Finally, when considering the combined cost of simultaneous trial incongruency and switch, a significant main effect of APOE4 status was reported (*F*(1, 56)=4,08, *p*=.048), driven by APOE4+ (*M*=.13, *SD=*.14) individuals showing greater cost than APOE33 counterparts (*M*=.08, *SD=*.13). In addition, the beneficial effect of levetiracetam (*M*=.09, *SD=*.12) on dual-cost relative to placebo (*M*=.13, *SD=*.11) trended towards significance (*F*(1, 56)=3.48, *p*=.067). The APOE4 status by drug condition interaction term was non-significant (*p*>.05). Effects of APOE4 gene dose on accuracy cost metrics are reported in Supplementary Materials.

**Table 2.** Descriptive statistics showing accuracy cost metrics by APOE4 status and Drug Condition. Values represent mean ± sd unless stated otherwise. * denotes significant differences by APOE4 status.

|  | Congruency Cost | Switch Cost | Dual cost |
| --- | --- | --- | --- |
| **APOE33** |  |  |  |
| Placebo | .06 ± .09 | .03 ± .04 | .10 ± .09 |
| **Levetiracetam** | .04 ± .05 | .02 ± .04 | .06 ± .08 |
| **APOE4+** |  |  |  |
| **Placebo** | .10 ± .09 | .04 ± .05 | .14 ± .13 |
| **Levetiracetam** | .09 ± .12 | .02 ± .05 | .11 ± .14 |

## Discussion

The present study tested whether low-dose levetiracetam enhanced cognitive performance in mid-age adults differentiated by APOE4 genetic risk, utilising a test of executive function anticipated to be sensitive to early genotype differences.

Current results show APOE4+ carriers exhibit poorer executive function in mid-life compared to APOE4 non-carriers, characterised by longer RTs and greater cost of increased inhibitory and dual (inhibition+switch) executive demands on performance accuracy. Treatment with low-dose levetiracetam was associated with improvements in executive performance, although the magnitude of benefit differed by genotype. The largest effects were observed in APOE33 individuals, who showed improvements in both RT and accuracy. In contrast, APOE4+ carriers demonstrated more selective benefits, limited to faster response times on low-demand congruent trials. Drug effects were further moderated by age and circulating biomarkers: cognitive benefits were greatest in younger participants and in those with lower levels of plasma indicators of neurodegeneration. Notably, by mid-life, APOE4+ carriers already showed significantly higher levels of amyloid-β, neurodegeneration (NfL) and mitochondrial dysfunction (GFAP) than their APOE33 counterparts. These underlying biological differences may partly explain why the cognitive effects of low-dose levetiracetam were attenuated in this higher-risk group.

APOE4+ status was associated with reduced executive function in mid-life, supporting prior research suggesting this cognitive domain is sensitive to emerging genotype differences[40,43,44,52,53 but see 54,55]. The experimental paradigm used here placed unpredictable, sometimes simultaneous demands on task-switching and inhibition; similar executive paradigms have discriminated individuals who subsequently convert to dementia up to 10 years pre-diagnosis[45], corroborating the task’s early sensitivity to AD risk. RTs were consistently slower in midlife APOE4+ individuals across all trial types, including those with low executive demand. This pattern may reflect domain-general cognitive slowing[39,53,56] potentially linked to genotype differences in integrity of the locus coeruleus (LC). The LC is vulnerable to early age-related changed and neurological stress including tau pathology[57,58]. Changes in noradrenergic signalling from the LC impacts both tonic cognitive arousal and executive processing in response to variable task demands[59,60]. One possibility is that slowing in mid-age APOE4+ may reflect accelerated ageing, including earlier pathology onset, in this critical brain region, having more widespread effects on fronto-parietal networks.

Notably, APOE4+ individuals in this study exhibited significantly higher levels of pathology, raising the possibility that observed genotype effects reflect a greater proportion of individuals in prodromal stages of Alzheimer’s disease. In cognitively healthy older adults, the detrimental impact of APOE4+ on attentional control has been shown to be mediated by amyloid pathology[61]. In contrast, during midlife, APOE4+ has been associated with working memory advantages, particularly under conditions of elevated amyloid-β[62]. However, the absence of a significant effect of amyloid-β on switch-inhibiton performance, as well as a lack of interaction with APOE4 status in the present study, suggests APOE4 may be affecting cognitive performance directly, as well as via its effects on enhanced pathology accumulation.

Low-dose levetiracetam was associated with acute benefits in executive function, aligning with previously reports this drug advantages executive function in healthy older adults[63,64] and AD populations with subclinical epileptiform activity[24,25]. Low-dose levetiracetam has also been associated with improved episodic memory[7,17] in individuals with MCI, and a reduction in entorhinal cortex atrophy in APOE4 non-carriers with MCI, strengthening the case for levetiracetam’s role as an early intervention for the treatment of neural hyperexcitability. In this study, beneficial effects of levetiracetam were greater at younger ages, in APOE33 individuals, and in individuals with lower-levels of pathology (as proxied through plasma). This may indicate the beneficial effects of levetiracetam on cognition are stronger in individuals with better neurological health.

Levetiracetam was hypothesised to confer greater benefit in APOE4 carriers, given evidence that lifespan patterns of neural hyperactivity in this group may drive pathological progression and cognitive decline (e.g., 32:34,37). However, in the present study, beneficial effects in APOE4+ individuals were restricted to trials with lower executive demand, in contrast to the broader cognitive benefits observed in APOE33 individuals. Prior work indicates that cognitively healthy midlife APOE4+ individuals exhibit elevated BOLD responses at lower working memory loads, but fail to appropriately scale activation with increasing task demands[55,65]. If low-dose levetiracetam exerts its effects by modulating functional hyperactivity, this may account for the selective benefits observed under lower cognitive load in APOE4+ individuals. The attenuated effects may also reflect the higher pathological burden observed in this group by midlife. Consistent with evidence of earlier pathological changes in APOE4 carriers[66], these findings underscore the importance of earlier intervention in this high-risk population, potentially before the emergence of cognitive and biological detriments. Future research may consider non-pharmacological interventions (e.g., neurofeedback[67]) as complementary, preventative approaches, theorised to also act on signatures of hyperactivity.

An 18-month intervention with AGB101, an extended-release low dose formulation of levetiracetam, reported a slowing of cognitive decline in APOE33, but not APOE4+, individuals with MCI[26]. As all participants exhibited significant amyloid pathology, differences in amyloid burden alone are unlikely to explain genotype-specific treatment effects. In addition, prior work suggests no marked genotype differences in BOLD activation at this stage of disease progression[68]. Levetiracetam primarily acts via binding to SV2A, a synaptic vesicle protein involved in vesicle fusion, recycling, and neurotransmitter release[69]. Reduced SV2A availability reported in cognitively healthy older APOE4+ individuals may therefore contribute to diminished treatment efficacy in this group[70]. Alternatively, increased damage to inhibitory interneurons in APOE4+ may impede restoration of excitation-inhibition balance in this higher risk group[40]. Emerging evidence indicates that levetiracetam can alter APP trafficking and reduce amyloidogenic processing at the presynaptic terminal[71]. These findings raise the possibility of levetiracetam’s disease-modifying effects if used earlier and longer-term – a critical avenue for future preventative research.

The current study is strengthened by its randomised, crossover, placebo-controlled design. The sample size, however, was defined by an *a priori* power calculation based on APOE4 differences in task-related fMRI BOLD response[33], in keeping with the larger research design, rather than potentially smaller genotype-effects on cognition[41,42]. Furthermore, exploratory outcomes underscore the need for more adequately powered studies to isolate whether the effects of levetiracetam differ by APOE4 gene dose, plus whether genotype differences are relate directly to differences in Alzheimer’s pathology (“healthy brain hypothesis”) or to inherent differences in levetiracetam’s mechanism of action in APOE4+ individuals. In line with past work [7,17,69], this study tested for an acute effect of low-dose levetiracetam on cognitive performance. Given that proposed mechanisms of action also implicate longer-term modulation of amyloid accumulation, future studies should examine the prophylactic potential of sustained treatment (e.g., ≥12 months) in midlife individuals at elevated risk, alongside any short-term benefits mediated by reductions in functional hyperactivity. Finally, the limited diversity of this proof-of-concept sample precluded examination of how sex and genetic ancestry may moderate treatment response, despite evidence these factors interact with APOE4 to influence cognition and brain function [52,72]; plus alter therapeutic efficacy[73] in AD.

In conclusion, current findings highlight midlife as a critical window for intervention in APOE4 carriers, during which emerging cognitive and biological changes may still be modifiable. While low-dose levetiracetam demonstrated acute cognitive benefits, particularly in individuals at lower risk of future AD, its effects in APOE4+ individuals were more limited, underscoring the need for earlier and potentially sustained intervention in this high-risk group. Given emerging evidence that levetiracetam may also influence synaptic and amyloid-related processes, longer-term administration warrants investigation as a preventative strategy. Targeting neural dysfunction prior to substantial pathological accumulation may be key to maximising therapeutic efficacy in APOE4+ individuals.

## Data Availability

Anonymised trial-level datasets are uploaded to the OSF (https://osf.io/2h3m9/files/rywtu).

https://osf.io/2h3m9/overview

## Acknowledgments

The authors thank Professor Sarah King (University of Sussex) for acting as the independent researcher responsible for randomisation and safety monitorying, plus Dr Prince Nwaubani and Dr Ed Caddye (Brighton & Sussex Medical School) for their support with blood collection. In addition, we acknowledge the Chalfont Centre for Epilepsy (UK) and Dementia Research Institute Biomarker Factory (University College London, UK), notably Dr Amanda Heslegrave, for the extraction of plasma-based metrics reported in this paper. The authors acknowledge the contribution of Dr Chris Hinds for providing the Mezurio smartphone app for compliance monitoring. This work was supported by funding from Alzheimer’s Society (AS-JF-19(a)-007) to C.L., and an Alzheimer’s Research UK South Coast Network pump priming grant to C.L., C.B., and J.R.

## References

1. Boxer, A. L., & Sperling, R. (2023). Accelerating Alzheimer’s therapeutic development: the past and future of clinical trials. Cell, 186(22), 4757–4772. 10.1016/j.cell.2023.09.023

2. Self, W. K., & Holtzman, D. M. (2023). Emerging diagnostics and therapeutics for Alzheimer disease. Nature medicine, 29(9), 2187–2199. 10.1038/s41591-023-02505-2

3. Chen, Y., Jin, H., Chen, J., Li, J., Găman, M. A., & Zou, Z. (2025). The multifaceted roles of apolipoprotein E4 in Alzheimer’s disease pathology and potential therapeutic strategies. Cell Death Discovery, 11(1), 312. 10.1038/s41420-025-02600-y

4. Steele, O. G., Stuart, A. C., Minkley, L., Shaw, K., Bonnar, O., Anderle, S., … & King, S. (2022). A multi-hit hypothesis for an APOE4-dependent pathophysiological state. European Journal of Neuroscience, 56(9), 5476–5515. 10.1111/ejn.15685

5. Sperling, R., Mormino, E., & Johnson, K. (2014). The evolution of preclinical Alzheimer’s disease: implications for prevention trials. Neuron, 84(3), 608–622. 10.1016/j.neuron.2014.10.038

6. Farrer, L. A., Cupples, L. A., Haines, J. L., Hyman, B., Kukull, W. A., Mayeux, R., … & Van Duijn, C. M. (1997). Effects of age, sex, and ethnicity on the association between apolipoprotein E genotype and Alzheimer disease: a meta-analysis. Jama, 278(16), 1349–1356. doi:10.1001/jama.1997.03550160069041

7. Bakker, A., Krauss, G. L., Albert, M. S., Speck, C. L., Jones, L. R., Stark, C. E., … & Gallagher, M. (2012). Reduction of hippocampal hyperactivity improves cognition in amnestic mild cognitive impairment. Neuron, 74(3), 467–474. 10.1016/j.neuron.2012.03.023

8. Yassa, M. A., Stark, S. M., Bakker, A., Albert, M. S., Gallagher, M., & Stark, C. E. (2010). High-resolution structural and functional MRI of hippocampal CA3 and dentate gyrus in patients with amnestic mild cognitive impairment. Neuroimage, 51(3), 1242–1252. 10.1016/j.neuroimage.2010.03.040

9. Berron, D., Cardenas-Blanco, A., Bittner, D., Metzger, C. D., Spottke, A., Heneka, M. T., … & Düzel, E. (2019). Higher CSF tau levels are related to hippocampal hyperactivity and object mnemonic discrimination in older adults. Journal of Neuroscience, 39(44), 8788–8797. 10.1523/JNEUROSCI.1279-19.2019

10. Busche MA, Konnerth A. Neuronal hyperactivity—A key defect in Alzheimer’s disease? BioEssays. 2015;37(6):624–632. doi:10.1002/bies.201500004

11. Huijbers W, Mormino EC, Schultz AP, et al. Amyloid-β deposition in mild cognitive impairment is associated with increased hippocampal activity, atrophy and clinical progression. Brain. 2015;138(Pt 4):1023–1035. doi:10.1093/brain/awv007

12. Miller SL, Fenstermacher E, Bates J, Blacker D, Sperling RA, Dickerson, BC. Hippocampal activation in adults with mild cognitive impairment predicts subsequent cognitive decline. J Neurol Neurosurg Psychiatry. 2008;79(6):630–635. doi:10.1136/jnnp.2007.124149

13. Leal, S. L., Landau, S. M., Bell, R. K., & Jagust, W. J. (2017). Hippocampal activation is associated with longitudinal amyloid accumulation and cognitive decline. Elife, 6, e22978. 10.7554/eLife.22978

14. Corriveau-Lecavalier, N., Adams, J. N., Fischer, L., Molloy, E. N., & Maass, A. (2024). Cerebral hyperactivation across the Alzheimer’s disease pathological cascade. Brain Communications, 6(6), fcae376. 10.1093/braincomms/fcae376

15. Haberman, R. P., Koh, M. T., & Gallagher, M. (2017). Heightened cortical excitability in aged rodents with memory impairment. Neurobiology of aging, 54, 144–151. 10.1016/j.neurobiolaging.2016.12.021

16. Sanchez, P. E., Zhu, L., Verret, L., Vossel, K. A., Orr, A. G., Cirrito, J. R., … & Mucke, L. (2012). Levetiracetam suppresses neuronal network dysfunction and reverses synaptic and cognitive deficits in an Alzheimer’s disease model. Proceedings of the National Academy of Sciences, 109(42), E2895–E2903. 10.1073/pnas.1121081109

17. Bakker, A., Albert, M. S., Krauss, G., Speck, C. L., & Gallagher, M. (2015). Response of the medial temporal lobe network in amnestic mild cognitive impairment to therapeutic intervention assessed by fMRI and memory task performance. NeuroImage: Clinical, 7, 688–698. 10.1016/j.nicl.2015.02.009

18. Koh, M., Haberman, R., Foti, S. et al. Treatment Strategies Targeting Excess Hippocampal Activity Benefit Aged Rats with Cognitive Impairment. Neuropsychopharmacol 35, 1016–1025 (2010). 10.1038/npp.2009.20

19. Devi, L., & Ohno, M. (2013). Effects of levetiracetam, an antiepileptic drug, on memory impairments associated with aging and Alzheimer’s disease in mice. Neurobiology of learning and memory, 102, 7–11. 10.1016/j.nlm.2013.02.001

20. Zheng, X. Y., Zhang, H. C., Lv, Y. D., Jin, F. Y., Wu, X. J., Zhu, J., & Ruan, Y. (2022). Levetiracetam alleviates cognitive decline in Alzheimer’s disease animal model by ameliorating the dysfunction of the neuronal network. Frontiers in aging neuroscience, 14, 888784. 10.3389/fnagi.2022.888784

21. Shi, J.-Q., Wang, B.-R., Tian, Y.-Y., Xu, J., Gao, L., Zhao, S.-L., Jiang, T., Xie, H.-G. and Zhang, Y.-D. (2013), Antiepileptics Topiramate and Levetiracetam Alleviate Behavioral Deficits and Reduce Neuropathology in APPswe/PS1dE9 Transgenic Mice. CNS Neurosci Ther, 19: 871–881. 10.1111/cns.12144

22. Lin, C. Y., Chang, M. C., & Jhou, H. J. (2024). Effect of levetiracetam on cognition: a systematic review and meta-analysis of double-blind randomized placebo-controlled trials. CNS drugs, 38(1), 1–14. 10.1007/s40263-023-01058-9

23. Faini, C., Sen, A., & Romoli, M. (2025). Effect of levetiracetam on cognition in patients with cognitive decline: A systematic review and meta-analysis of randomized controlled trials. Epilepsia Open. 10.1002/epi4.70091

24. Vossel, K., Ranasinghe, K. G., Beagle, A. J., La, A., Pook, K. A., Castro, M., … & Kirsch, H. E. (2021). Effect of levetiracetam on cognition in patients with Alzheimer disease with and without epileptiform activity: a randomized clinical trial. JAMA neurology, 78(11), 1345–1354. doi:10.1001/jamaneurol.2021.3310

25. Musaeus, C. S., Nilsson, C., Cooper, C., Kramberger, M. G., Verdelho, A., Stefanova, E., … & Frederiksen, K. S. (2021). Pharmacological medical treatment of epilepsy in patients with dementia: A systematic review. Current Alzheimer Research, 18(9), 689–694. 10.2174/1567205018666211126121529

26. Mohs R, Bakker A, Rosenzweig-Lipson S, et al. The HOPE4MCI study: A randomized double-blind assessment of AGB101 for the treatment of MCI due to AD. Alzheimer’s Dement. 2024;10::e12446. 10.1002/trc2.12446

27. Bakker A, Rani N, Mohs R, Gallagher M. The HOPE4MCI study: AGB101 treatment slows progression of entorhinal cortex atrophy in APOE ε4 non-carriers with mild cognitive impairment due to Alzheimer’s disease. Alzheimer’s Dement. 2024;e70004. 10.1002/trc2.70004

28. Cummings, J., Apostolova, L., Rabinovici, G. D., Atri, A., Aisen, P., Greenberg, S., … & Salloway, S. (2023). Lecanemab: appropriate use recommendations. The journal of prevention of Alzheimer’s disease, 10(3), 362–377.

29. Lee, H., Stirnberg, R., Wu, S., Wang, X., Stöcker, T., Jung, S., … & Axmacher, N. (2020). Genetic Alzheimer’s disease risk affects the neural mechanisms of pattern separation in hippocampal subfields. Current Biology, 30(21), 4201–4212. 10.14283/jpad.2023.30

30. Filippini, N., MacIntosh, B. J., Hough, M. G., Goodwin, G. M., Frisoni, G. B., Smith, S. M., … & Mackay, C. E. (2009). Distinct patterns of brain activity in young carriers of the APOE-ε4 allele. Proceedings of the National Academy of Sciences, 106(17), 7209–7214. 10.1073/pnas.0811879106

31. Sinha, N., Berg, C. N., Tustison, N. J., Shaw, A., Hill, D., Yassa, M. A., & Gluck, M. A. (2018). APOE ε4 status in healthy older African Americans is associated with deficits in pattern separation and hippocampal hyperactivation. Neurobiology of aging, 69, 221–229. 10.1016/j.neurobiolaging.2018.05.023

32. Wishart, H. A., Saykin, A. J., Rabin, L. A., Santulli, R. B., Flashman, L. A., Guerin, S. J., … & McAllister, T. W. (2006). Increased brain activation during working memory in cognitively intact adults with the APOE ε4 allele. American Journal of Psychiatry, 163(9), 1603–1610. 10.1176/ajp.2006.163.9.160

33. Scheller, E., Schumacher, L. V., Peter, J., Lahr, J., Wehrle, J., Kaller, C. P., … & Klöppel, S. (2018). Brain aging and APOE ε4 interact to reveal potential neuronal compensation in healthy older adults. Frontiers in aging neuroscience, 10, 74. 10.3389/fnagi.2018.00074

34. Rabipour, S., Rajagopal, S., Yu, E., Pasvanis, S., Lafaille-Magnan, M. E., Breitner, J., … & Rajah, M. N. (2020). APOE4 status is related to differences in memory-related brain function in asymptomatic older adults with family history of Alzheimer’s disease: baseline analysis of the PREVENT-AD task functional MRI dataset. Journal of Alzheimer’s Disease, 76(1), 97–119. 10.3233/JAD-191292

35. Henson, R. N., Suri, S., Knights, E., Rowe, J. B., Kievit, R. A., Lyall, D. M., … & Fisher, S. E. (2020). Effect of apolipoprotein E polymorphism on cognition and brain in the Cambridge Centre for Ageing and Neuroscience cohort. Brain and Neuroscience Advances, 4, 2398212820961704. 10.1177/2398212820961704

36. Raykov PP, Daly J, Fisher SE, Eising E, Geerligs L, Bird CM. No effect of apolipoprotein E polymorphism on MRI brain activity during movie watching. Brain and Neuroscience Advances. 2025;9. doi:10.1177/23982128251314577

37. Tabuena, D. R., Jang, S. S., Grone, B., Yip, O., Aery Jones, E. A., Blumenfeld, J., … & Zilberter, M. (2026). Neuronal APOE4-induced early hippocampal network hyperexcitability in Alzheimer’s disease pathogenesis. Nature Aging, 1–19. 10.1038/s43587-026-01096-0

38. Lancaster, C., Tabet, N., & Rusted, J. (2017). The elusive nature of APOE ε4 in mid-adulthood: Understanding the cognitive profile. Journal of the International Neuropsychological Society, 23(3), 239–253. doi:10.1017/S1355617716000990

39. Lancaster, C., Berens, S., Daly, J., Rusted, J., & Bird, C. M. (2025). Perceptual discrimination of complex objects: Apolipoprotein E e4 gene-dose effects in mid-life. Alzheimer’s & Dementia, 21(6), e70246. 10.1002/alz.70246

40. Lancaster, C., Tabet, N., & Rusted, J. (2016). The APOE paradox: do attentional control differences in mid-adulthood reflect risk of late-life cognitive decline. Neurobiology of aging, 48, 114–121. 10.1016/j.neurobiolaging.2016.08.015

41. Lancaster, C., McDaniel, M. A., Tabet, N., & Rusted, J. (2020). Prospective memory: Age related change is influenced by APOE genotype. Aging, Neuropsychology, and Cognition, 27(5), 710–728. doi:10.1080/13825585.2019.1671305

42. Goveas, J.S., Xie, C., Chen, G., Li, W., Ward, B.D., Franczak, M.B., & Li, S.-J. (2013). Functional network endophenotypes unravel the effects of apolipoprotein E epsilon 4 in middle-aged adults. PloS One, 8(2), e55902. doi: 10.1371/journal.pone.0055902

43. Marioni, R.E., Campbell, A., Scotland, G., Hayward, C., Porteous, D.J., & Deary, I.J.(2016). Differential effects of the APOE ε4 allele on different domains of cognitive ability across the life-course. European Journal of Human Genetics, 24, 919–923. doi: 10.1038/ejhg.2015.210

44. Velichkovsky, B.B., Roschina, I.F., & Selezneva, N.D. (2015). Cognitive control and memory in healthy ApoE-ε4 carriers with a family history of Alzheimer’s disease. Psychology in Russia: State of the Art, 8(1), 4–13. doi: 10.11621/pir.2015.0101

45. Balota, D. A., Tse, C. S., Hutchison, K. A., Spieler, D. H., Duchek, J. M., & Morris, J. C. (2010). Predicting conversion to dementia of the Alzheimer’s type in a healthy control sample: The power of errors in stroop color naming. Psychology and aging, 25(1), 208. doi:10.1037/a0017474.

46. Hutchison, K. A., Balota, D. A., & Ducheck, J. M. (2010). The utility of Stroop task switching as a marker for early-stage Alzheimer’s disease. Psychology and aging, 25(3), 545. doi:10.1037/a0018498.

47. Lancaster, C., Koychev, I., Blane, J., Chinner, A., Chatham, C., Taylor, K., & Hinds, C. (2020). Gallery Game: Smartphone-based assessment of long-term memory in adults at risk of Alzheimer’s disease. Journal of clinical and experimental neuropsychology, 42(4), 329–343. 10.1080/13803395.2020.1714551

48. Lancaster, C., Koychev, I., Blane, J., Chinner, A., Wolters, L., & Hinds, C. (2020). Evaluating the feasibility of frequent cognitive assessment using the Mezurio smartphone app: observational and interview study in adults with elevated dementia risk. JMIR mHealth and uHealth, 8(4), e16142. 10.2196/16142

49. Belloy, M. E., Andrews, S. J., Le Guen, Y., Cuccaro, M., Farrer, L. A., Napolioni, V., & Greicius, M. D. (2023). APOE genotype and Alzheimer disease risk across age, sex, and population ancestry. JAMA neurology, 80(12), 1284–1294. doi:10.1001/jamaneurol.2023.3599

50. Suri, S., Heise, V., Trachtenberg, A. J., & Mackay, C. E. (2013). The forgotten APOE allele: a review of the evidence and suggested mechanisms for the protective effect of APOE ɛ2. Neuroscience & Biobehavioral Reviews, 37(10), 2878–2886. 10.1016/j.neubiorev.2013.10.010.

51. Nelson, H. E., & Willison, J. (1991). National adult reading test (NART) (pp. 1–26). Windsor: Nfer-Nelson.

52. Greenwood, P. M., Lambert, C., Sunderland, T., & Parasuraman, R. (2005). Effects of apolipoprotein E genotype on spatial attention, working memory, and their interaction in healthy, middle-aged adults: results From the National Institute of Mental Health’s BIOCARD study. Neuropsychology, 19(2), 199.

53. Evans, S., Dowell, N. G., Tabet, N., Tofts, P. S., King, S. L., & Rusted, J. M. (2014). Cognitive and neural signatures of the APOE E4 allele in mid-aged adults. Neurobiology of aging, 35(7), 1615–1623. 10.1016/j.neurobiolaging.2014.01.145

54. Trachtenberg, A. J., Filippini, N., Cheeseman, J., Duff, E. P., Neville, M. J., Ebmeier, K. P., … & Mackay, C. E. (2012). The effects of APOE on brain activity do not simply reflect the risk of Alzheimer’s disease. Neurobiology of aging, 33(3), 618–e1. 10.1016/j.neurobiolaging.2010.11.011

55. Chen, C. J., Chen, C. C., Wu, D., Chi, N. F., Chen, P. C., Liao, Y. P., … & Hu, C. J. (2013). Effects of the apolipoprotein E ε4 allele on functional MRI during n-back working memory tasks in healthy middle-aged adults. American Journal of Neuroradiology, 34(6), 1197–1202.

56. Luo, X., Jiaerken, Y., Yu, X., Huang, P., Qiu, T., Jia, Y., … & Alzheimer’s Disease Neuroimaging Initiative (ADNI). (2017). Affect of APOE on information processing speed in non-demented elderly population: a preliminary structural MRI study. Brain Imaging and Behavior, 11(4), 977–985.

57. Bueichekú, E., Diez, I., Kim, C. M., Becker, J. A., Koops, E. A., Kwong, K., … & Jacobs, H. I. (2024). Spatiotemporal patterns of locus coeruleus integrity predict cortical tau and cognition. Nature aging, 4(5), 625–637. 10.1038/s43587-024-00626-y

58. Dahl, M. J., Mather, M., & Werkle-Bergner, M. (2022). Noradrenergic modulation of rhythmic neural activity shapes selective attention. Trends in cognitive sciences, 26(1), 38–52. 10.1016/j.tics.2021.10.009

59. Unsworth, N., & Robison, M. K. (2017). A locus coeruleus-norepinephrine account of individual differences in working memory capacity and attention control. Psychonomic bulletin & review, 24(4), 1282–1311. 10.3758/s13423-016-1220-5

60. Robison, M. K., Ralph, K. J., Gondoli, D. M., Torres, A., Campbell, S., Brewer, G. A., & Gibson, B. S. (2023). Testing locus coeruleus-norepinephrine accounts of working memory, attention control, and fluid intelligence. Cognitive, Affective, & Behavioral Neuroscience, 23(4), 1014–1058. 10.3758/s13415-023-01096-2

61. Aschenbrenner, A. J., Balota, D. A., Tse, C. S., Fagan, A. M., Holtzman, D. M., Benzinger, T. L., & Morris, J. C. (2015). Alzheimer disease biomarkers, attentional control, and semantic memory retrieval: Synergistic and mediational effects of biomarkers on a sensitive cognitive measure in non-demented older adults. Neuropsychology, 29(3), 368. 10.1037/neu0000133

62. Lu, K., Nicholas, J. M., Pertzov, Y., Grogan, J., Husain, M., Pavisic, I. M., … & Crutch, S. J. (2021). Dissociable effects of APOE ε4 and β-amyloid pathology on visual working memory. Nature Aging, 1(11), 1002–1009. 10.1038/s43587-021-00117-4

63. Schoenberg, M. R., Rum, R. S., Osborn, K. E., & Werz, M. A. (2017). A randomized, double-blind, placebo-controlled crossover study of the effects of levetiracetam on cognition, mood, and balance in healthy older adults. Epilepsia, 58(9), 1566–1574. 10.1111/epi.13849

64. Magalhães, J. C., Gongora, M., Vicente, R., Bittencourt, J., Tanaka, G., Velasques, B., … & Ribeiro, P. (2015). The influence of levetiracetam in cognitive performance in healthy individuals: neuropsychological, behavioral and electrophysiological approach. Clinical Psychopharmacology and Neuroscience, 13(1), 83. doi: 10.9758/cpn.2015.13.1.83

65. Yan, F. X., Wu, C. W., Chao, Y. P., Chen, C. J., & Tseng, Y. C. (2015). APOE-ε4 allele altered the rest-stimulus interactions in healthy middle-aged adults. PLoS One, 10(6), e0128442. 10.1371/journal.pone.0128442

66. Mishra, S., Blazey, T. M., Holtzman, D. M., Cruchaga, C., Su, Y., Morris, J. C., … & Gordon, B. A. (2018). Longitudinal brain imaging in preclinical Alzheimer disease: impact of APOE ε4 genotype. Brain, 141(6), 1828–1839. 10.1093/brain/awy103

67. Klink, K., Lysser, Y., Wunderlin, M., Radojewski, P., Teunissen, C., Orth, M., & Peter, J. (2025). Downregulation of hippocampal activity improves memory performance in individuals at risk of Alzheimer’s disease. medRxiv, 2025–10.

68. Tran, T. T., Speck, C. L., Pisupati, A., Gallagher, M., & Bakker, A. (2017). Increased hippocampal activation in ApoE-4 carriers and non-carriers with amnestic mild cognitive impairment. NeuroImage: Clinical, 13, 237–245. 10.1016/j.nicl.2016.12.002

69. Nussbaumer, J., Barve, A., Zufferey, V., Espourteille, J., Kirabali, T., Konietzko, U., … & Ni, R. (2025). Reduced synaptic vesicle protein 2A in extracellular vesicles and brains of Alzheimer’s disease: associations with Aβ, tau, synaptic proteins and APOE ε4. Translational Neurodegeneration, 14(1), 48. 10.1186/s40035-025-00508-2

70. Snellman, A., Tuisku, J., Koivumäki, M., Wahlroos, S., Aarnio, R., Rajander, J., … & Rinne, J. O. (2024). SV2A PET shows hippocampal synaptic loss in cognitively unimpaired APOE ε4/ε4 homozygotes. Alzheimer’s & Dementia, 20(12), 8802–8813. 10.1002/alz.14327

71. Rao, N. R., Santiago-Marrero, I., DeGulis, O., Nomura, T., Goyal, K., Lee, S., … & Savas, J. N. (2026). Levetiracetam prevents Aβ production through SV2a-dependent modulation of APP processing in Alzheimer’s disease models. Science Translational Medicine, 18(836), eadp3984. DOI: 10.1126/scitranslmed.adp3984

72. Altmann, A., Tian, L., Henderson, V. W., Greicius, M. D., & Alzheimer’s Disease Neuroimaging Initiative Investigators. (2014). Sex modifies the APOE-related risk of developing Alzheimer disease. Annals of neurology, 75(4), 563–573. 10.1002/ana.24135

73. Van Dyck, C. H., Swanson, C. J., Aisen, P., Bateman, R. J., Chen, C., Gee, M., … & Iwatsubo, T. (2023). Lecanemab in early Alzheimer’s disease. New England Journal of Medicine, 388(1), 9–21. 10.1056/NEJMoa2212948

