## Supplementary Materials for "APOE-specific Cognitive Effects of Levetiracetam in Mid-Age Adults"

**Supplementary Table 1.** Descriptive statistics showing proportion accuracy (Acc) and response time (RT(ms) correct trials only) on each trial type (congruent, neutral, incongruent; no-switch (No-S), switch (S)), grouped by APOE4 status and drug condition.

| | | APOE33 | | APOE34 | |
|---|---|---|---|---|---|
| **Trial Type** | **Outcome** | **Placebo** | **Levetiracetam** | **Placebo** | **Levetiracetam** |
| Congruent | | | | | |
| | Acc | .98 ± .07 | .98 ± .05 | .99 ± .04 | .97 ± .13 |
| No-S | RT (ms) | 689 ± 134 | 674 ± 159 | 807 ± 237 | 721 ± 132 |
| | Acc | .96 ± .08 | .98 ± .04 | .99 ± .05 | .97 ± .09 |
| S | RT (ms) | 840 ± 281 | 750 ± 119 | 933 ± 264 | 832 ± 178 |
| Neutral | | | | | |
| | Acc | .98 + .07 | 1.00 ± .00 | .98 ± .09 | .98 ± .10 |
| No-S | RT (ms) | 753 ± 227 | 666 ± 108 | 758 ± 182 | 753 ± 249 |
| | Acc | .99 ± .04 | .98 ± .06 | .96 ± .08 | .98 ± .11 |
| S | RT (ms) | 861 ± 252 | 742 ± 143 | 923 ± 343 | 829 ± 131 |
| Incongruent | | | | | |
| | Acc | .93 ± .11 | .96 ± .06 | .92 ± .12 | .91 ± .12 |
| No-S | RT (ms) | 838 ± 241 | 755 ± 123 | 914 ± 190 | 848 ± 179 |
| | Acc | .87 ± .12 | .92 ± .08 | .83 ± .14 | .87 ± .12 |
| S | RT (ms) | 948 ± 250 | 853 ± 248 | 981 ± 280 | 963 ± 165 |

**Effect of APOE4 status and low-dose Levetiracetam on performance on neutral trials.**

Response times to neutral trials (767ms ± 398ms) are faster than for congruent (790ms ± 424ms) trials; this does not suggest response speed was facilitated by congruent direction-location cues. Consistent with Model 1, main effects of trial switch ($t(2067)=6.66$, $p<.001$), $z$-standardised age ($t(2067)=2.48$, $p=.013$), and drug condition ($t(2067)=-2.45$, $p=.014$) were reported on RTs for correct neutral trials, with slower RTs estimated for switch trials, with increasing participant age, and under placebo. In addition, significant interactions between Condition and APOE4 ($t(2067)=2.38$, $p=.017$), and z-standardised age and APOE4 ($t(2067)=1.987$, $p=.047$) were reported. Beneficial effects of levetiracetam on trial RT were only significant in APOE33 individuals ($p<.001$). In addition, the effect of age was only significant in APOE4+ individuals ($p=.001$), driven by these individuals being faster below mean sample age, and slower thereafter.

There was no evidence that performance accuracy (Model 2) was facilitated on congruent (*M*=.98 ± .08) relative to neutral (*M*=.98 ± .08) trials. Main effects of trial switch, condition, and APOE4 status on neutral trial accuracy were non-significant ($p>.05$), as were all interaction terms ($p>.05$). Performance on neutral trials, however, was at ceiling.

**Sex as a predictor of switch-inhibition performance.**

Adding Sex as a covariate into Model 1 (Estimated RT) did not significantly improve model fit. However, there was a main effect of Sex ($t(5939)=3.25$, $p=.001$), driven by females responding slower than males. All other main effects and interaction terms remained unchanged. Given the smaller number of males in this sample, interactions between Sex, APOE4, and drug condition were not explored further.

The addition of Sex (male=0, female=1) as a binary predictor did not improve fit of Model 2 – probability of correct response. There was a non-significant main effect of Sex ($p>.05$).

**APOE4 gene dose as a predictor of switch-inhibition performance.**

There was a significant difference in age between the three genotype groups, $\chi^2(2)=6.96$, $p=.031$, driven by the APOE44 individuals being older. Group differences in all other demographic characteristics and plasma-biomarkers were non-significant (Supplementary Table 2).

**Supplementary Table 2.** Characteristics of mid-age participants broken down by APOE4 gene dose

| | **APOE33** | **APOE34** | **APOE44** |
|---|---|---|---|
| *n* | 27 | 23 | 8 |
| Age (Years) | 54.11 ± 4.91* | 55.57 ± 5.39* | 59.25 ± 2.60* |
| Sex (%F) | 70.37 | 82.61 | 62.5 |
| Education (Years) | 17.37 ± 3.31 | 18.13 ± 3.81 | 18.63 ± 4.78 |
| Full-scale Premorbid IQ | 119.55 ± 4.09 | 118.36 ± 4.77 | 115.55 ± 3.23 |
| MOCA (/30) | 27.81 ± 1.73 | 27.43 ± 1.70 | 26.75 ± 2.25 |
| Plasma levetiracetam concentration(mg/L) | 3.53 ± .84 | 3.37 ± 1.07 | 2.51 ± .85 |
| Aβ42:40 (pg/ml) | .07 ± .01 | .06 ± .01 | .06 ± .01 |
| p-tau181 (pg/ml) | 20.16 ± 8.34 | 24.90 ± 16.09 | 21.52 ± 7.68 |
| GFAP (pg/ml) | 74.70 ± 23.24 | 113.80 ± 73.13 | 144.41 ± 74.29 |
| NfL (pg/ml) | 8.68 ± 2.46 | 13.27 ± 6.24 | 12.89 ± 2.56 |

**Effect of APOE e4 gene dose and Levetiracetam on RT**

Replacing APOE4 status with APOE4 gene dose (linear) does not substantially affect model fit (AIC=83330). Model estimates are shown in Supplementary Figure 1. The main effect of APOE4 gene dose is non-significant ($t(5940)=.857$, $p=.391$), however all other main effects (drug condition, age, trial congruency, trial switch), plus the condition x age interaction term, remain significant ($p<.05$) as per the main analyses.

**Supplementary Figure 1.** *Frequentist model estimates of response time (RT) for correct trials, separated by Apolipoprotein E e4 gene dose and drug condition*

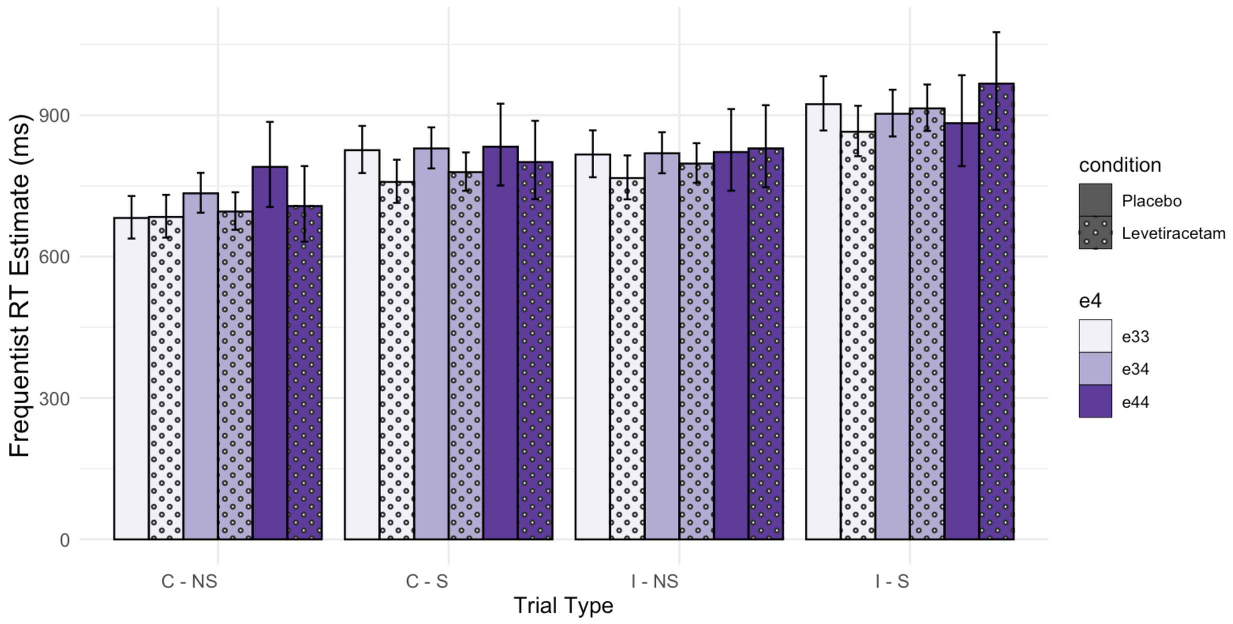


***Note.*** Trials are grouped as by congruency (congruent (C), incongruent (I)) and switch (no-switch (NS), switch (S)). Estimates are shown at mean sample age.

The following 3-way interaction terms are significant: 1) drug condition x dose x trial congruency ($t(5940)=-2.42$, $p=.016$); 2) drug condition x dose x trial switch ($t(5940)=1.98$, $p=.048$). The effect of APOE4 gene dose is not significant under any drug condition for both congruent and incongruent trials ($p>.006$). As in the main analyses, there is a significant, beneficial effect of levetiracetam on response speed in APOE33 individuals for incongruent trials ($p<.001$), with the positive effect only trending towards significance for congruent trials ($p=.045$) following correction for multiple comparisons ($\alpha=.003$). Levetiracetam selectively advantages performance in APOE34 individuals for congruent trials ($p<.001$), with beneficial effect of drug only trending towards significance for APOE44 individuals on this easier task condition =.031).

Likewise, the effect of APOE4 gene dose is not significant under any drug condition for both switch and non-switch trials ($p > .006$). A significant beneficial effect of levetiracetam is only reported on switch trials in APOE33 carriers ($p < .001$), however, drug effects trend towards significance for APOE34 individuals on no-switch trials ($p = .024$).

**Effect of APOE e4 gene dose and Levetiracetam on Accuracy**

Model 2 was re-run with dose included as a linear predictor, with a marginal improvement to model fit (AIC=2625, BIC = 2732). As in the main analyses, there was a significant effect of trial congruency ($t(6368) = -8.71$, $p < .001$) and trial switch ($t(6368) = -3.05$, $p = .002$) on probability of correct response, however, the main effects of drug condition and APOE4 gene dose were non-significant ($p > .05$). As before there was a significant condition by switch interaction, ($t(6368) = -2.09$, $p = .038$), plus interactions between APOE4 gene dose and trial congruency ($t(6368) = -3.39$, $p < .001$), and APOE4 gene dose and drug condition ($t(6368) = -3.24$, $p = .001$).

Post-hoc comparisons indicate there is a significant detrimental effect of gene-dose on accuracy for incongruent ($p = .043$, $\alpha = .010$) but not congruent trials ($p = .153$). All APOE4 gene dose groups showed a significant cost of trial incongruency, with the largest effect seen in APOE34 carriers (z-ratio=10.04), relative to APOE44 (z-ratio=7.07) and APOE333 (z-ratio=5.60) individuals.

The linear effect of APOE4 gene dose was non-significant under placebo and levetiracetam ($p > .010$). The positive effect of levetiracetam on response accuracy in APOE33 only approached significance ($p = .011$, $\alpha = .010$), however, there was a significant detrimental effect of levetiracetam on estimated probability of correct response in APOE44 individuals ($p = .006$). Model estimates are shown in Supplementary Figure 2.

**Supplementary Figure 2.** *Frequentist model estimates of probability of correct response, separated by Apolipoprotein E e4 gene dose and drug condition.*

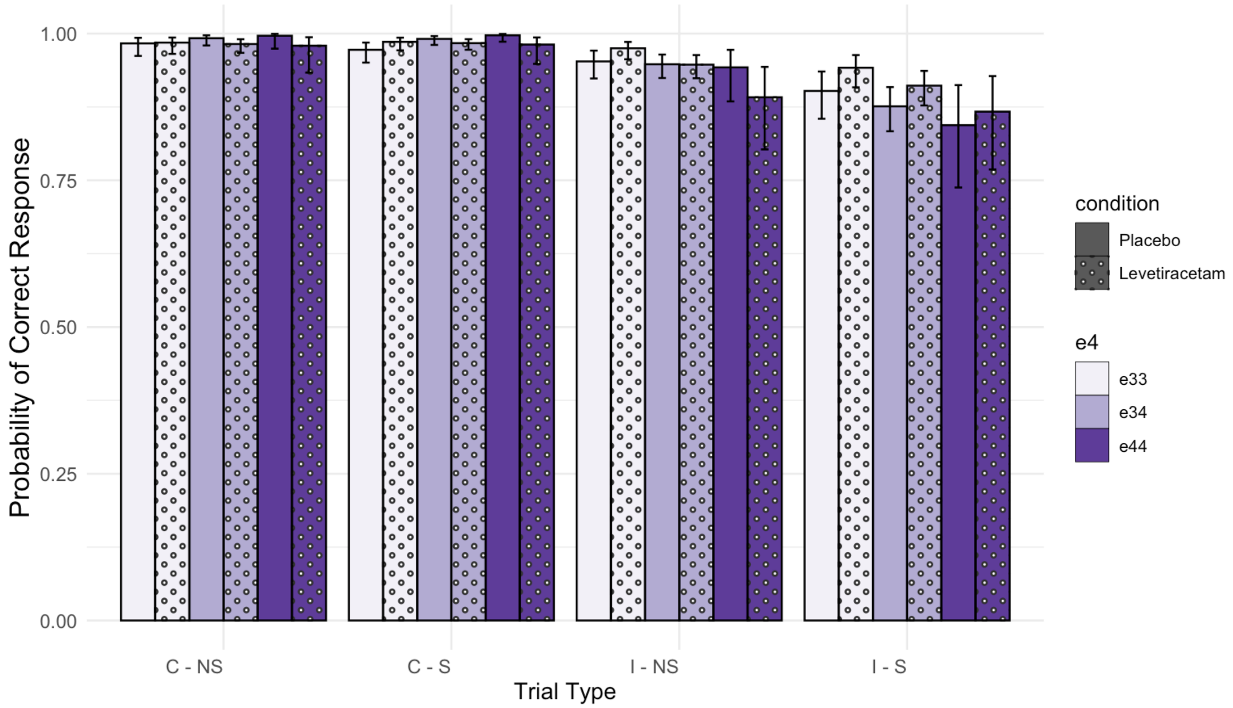


***Note.*** Trials are grouped as by congruency (congruent (C), incongruent (I)) and switch (no-switch (NS), switch (S)). Estimates are shown at mean sample age.

#### Accuracy Cost

There is significant effect of APOE4 gene dose on cost of trial incongruency ($F(2, 55)=3.62$, $p=.031$) with APOE44 individuals showing a significantly greater decline in accuracy ($M=.12$, $SD=.11$) than the APOE33 control group ($M=.05$, $SD=.07$; $p=.007$; $\alpha=.017$), but not APOE34 carriers ($M=.08$, $SD=.10$, $p>.017$). In addition, whilst the main effect of drug condition is non-significant ($p>.05$), there is a significant APOE4 gene dose x condition interaction ($F(2, 55)=3.62$, $p=.031$). Post-hoc comparisons ($\alpha=.008$) report significant group differences under levetiracetam only, driven by APOE44 individuals showing significantly greater congruency cost than APOE33 individuals ($p<.001$) and APOE34 individuals ($p=.005$). The effect of drug condition on congruency cost to task accuracy is non-significant in all 3 groups ($p>.008$).

There is a significant effect of drug condition on switch cost to trial accuracy ($F(2, 55)=4.77$, $p=.031$), driven by their being a larger cost to performance under placebo ($M=.03$, $SD=.04$) than levetiracetam ($M=.02$, $SD=.04$). The main effect of APOE4 gene dose and dose by condition interaction were both non-significant ($p>.05$).

In addition, the main effect of APOE4 gene dose, drug condition, and the APOE4 gene dose x drug condition interaction on the cost of dual-executive load to task accuracy was non-significant ($p$>.05). Descriptive statistics of task performance are shown in Supplementary Table 1.

### Inclusion of MoCA scores as an additional predictor of inhibition-switch performance

### RT.

Inclusion of MoCA as a standardised covariate marginally improved fit of Model 1 (AIC=83328). Lower scores on the MoCA predicted significantly longer RTs on the inhibition-switch task, $t$(5939)=-4.73, $p$<.001. All main effects and interaction terms reported in the main analyses remained unchanged except the main effect of APOE4 status now only approached significance, $t$(5939)=1.71, $p$=.087.

#### Accuracy.

Lower scores on the MoCA, entered as standardised covariate, significantly negatively predicted probability of a correct-response on the inhibition-switch task, $t$(6367)=2.72, $p$=.007. Following inclusion of this covariate, all main effects and interaction terms remained qualitatively the same.
